# Bayesian workflow for time-varying transmission in stratified compartmental infectious disease transmission models

**DOI:** 10.1101/2023.10.09.23296742

**Authors:** Judith A. Bouman, Anthony Hauser, Simon L. Grimm, Martin Wohlfender, Samir Bhatt, Elizaveta Semenova, Andrew Gelman, Christian L. Althaus, Julien Riou

## Abstract

Compartmental models that describe infectious disease transmission across subpopulations are central for assessing the impact of non-pharmaceutical interventions, behavioral changes and seasonal effects on the spread of respiratory infections. We present a Bayesian workflow for such models, including four features: (1) an adjustment for incomplete case ascertainment, (2) an adequate sampling distribution of laboratory-confirmed cases, (3) a flexible, time-varying transmission rate, and (4) a stratification by age group. We benchmarked the performance of various implementations of two of these features (2 and 3). For the second feature, we used SARS-CoV-2 data from the canton of Geneva (Switzerland) and found that a quasi-Poisson distribution is the most suitable sampling distribution for describing the overdispersion in the observed laboratory-confirmed cases. For the third feature, we implemented three methods: Brownian motion, B-splines, and approximate Gaussian processes (aGP). We compared their performance in terms of the number of effective samples per second, and the error and sharpness in estimating the time-varying transmission rate over a selection of ordinary differential equation solvers and tuning parameters, using simulated seroprevalence and laboratory-confirmed case data. Even though all methods could recover the time-varying dynamics in the transmission rate accurately, we found that B-splines perform up to four and ten times faster than Brownian motion and aGPs, respectively. We validated the B-spline model with simulated age-stratified data. We applied this model to 2020 laboratory-confirmed SARS-CoV-2 cases and two seroprevalence studies from the canton of Geneva. This resulted in detailed estimates of the transmission rate over time and the case ascertainment. Our results illustrate the potential of the presented workflow including stratified transmission to estimate age-specific epidemiological parameters. The workflow is freely available in the R package HETTMO, and can be easily adapted and applied to other surveillance data.

**Author summary:** Mathematical models are a central tool for understanding the spread of infectious diseases. These models can be fitted to surveillance data such as the number of laboratory-confirmed cases and seroprevalence over time. To provide insightful information for managing an epidemic, the models require several crucial features. In our study, we compare the performance of several implementations of two such features. First, we find that a quasi-Poisson distribution describes best how the number of laboratory-confirmed cases of SARS-CoV-2 from the canton of Geneva (Switzerland) are sampled from the total incidence of the infection. Second, we conclude that a B-spline based implementation of time-variation in the transmission rate performs better than a Brownian motion or approximate Gaussian processes based model. Moreover, we confirm that the B-spline based model can recover time-varying transmission also in an age-stratified population. This structural comparison of methods results in a Bayesian workflow. Such a comprehensive workflow is crucial to move the field of mathematical modeling for infectious disease dynamics forward and make methods widely applicable.

## Introduction

Epidemic theory provides mathematical expressions for biological concepts that are fundamental to understanding the spread of infectious diseases, such as contagion, incubation and immunity. Compartmental models based on ordinary differential equations (ODEs) implement these concepts within a unified, manageable framework, and have taken a central position in the field of infectious disease modeling. While initially used to formalize and develop theoretical notions such as reproductive numbers or immunity thresholds [1], or to simulate epidemics under specific constraints [2], [3], compartmental transmission models have been increasingly applied to practical questions about infectious disease transmission, especially during the SARS-CoV-2 pandemic [4]–[6]. These applications often rely upon fitting custom-made models to surveillance data such as counts of laboratory-confirmed cases, and use various methods of statistical inference. Among these, Bayesian inference with Markov chain Monte Carlo (MCMC) is gaining ground, fueled by improvements in computing power and sampling algorithms [7], and by efficient software implementations [8]–[10]. This approach offers many advantages, including parameter inference, full propagation of uncertainty, principled integration of prior knowledge and high flexibility in model specification [11]. Still, even the most basic situations require models of relatively high complexity, with many options available for each model feature, and difficulties of implementation and computational inefficiency limit the widespread adoption of these tools. Therefore, there is a growing demand for readily available Bayesian workflows, which have been through the process of model building, validation and comparison of different models [12].

We identified four essential features for a Bayesian workflow aimed at studying the transmission of SARS-CoV-2 (or other respiratory viruses) in a population over a relatively short time period based on counts of laboratory-confirmed cases: (1) an adjustment for incomplete case ascertainment, (2) an adequate sampling distribution of laboratory-confirmed cases, (3) a flexible, time-varying transmission rate, and (4) a stratification by age group. First, incomplete and unrepresentative ascertainment plays a key role in the generation of surveillance data. Indeed, laboratory-confirmed cases are only an unrepresentative subset of the actual population of newly infected individuals, that is highly dependent on testing activity (how many tests are performed) and targeting (which part of the population is prioritized for or has access to testing), both of which can vary over time [13]–[15]. The identification of the ascertainment rate, however, requires additional data such as point estimates of population seroprevalence [11]. Second, the sampling distribution must be suitable to generate counts of laboratory-confirmed cases. Common options include Poisson, quasi-Poisson and negative binomial distributions, but no systematic comparison in this context has been conducted to date [16], [17]. The third feature, flexible time-varying transmission, is critical, as it models the variations in transmission caused by drivers such as non-pharmaceutical interventions (NPIs), alterations of behaviors, and environmental determinants. These drivers can impact both components of the transmission rate: the rate of contact between individuals (e.g., mandatory work from home) and the probability of transmission upon contact (e.g., mandatory face masks). As these factors may vary over time, any model aimed at disentangling and understanding the drivers of SARS-CoV-2 transmission must incorporate a time-varying transmission rate. Several approaches have been proposed using predefined functional shapes [18]–[21] or more flexible approaches based on step functions [5], cubic splines [22]–[24] or Brownian motion [25]–[27]. A systematic comparison of these methods in the context of compartmental transmission models is currently lacking. Fourth, the stratification by age group is now considered standard practice in transmission models of respiratory infections [28]. Indeed, age influences every step of the infection course of SARS-CoV-2 and other respiratory viruses including contact patterns, adherence to NPIs, probability of testing and probability of severe outcome [27], [29]–[31]. While other individual factors like gender and socio-economic position may certainly influence transmission [32], age is generally considered as the most important, justifying this first choice for stratification.

In this work, we present a Bayesian workflow for a compartmental transmission model to analyze the transmission of respiratory viruses that includes these four essential features. To this aim, we assess the statistical accuracy and computational efficiency of several versions of the model, including three sampling distributions (Poisson, quasi-Poisson, and negative binomial) and three methods for implementing a time-varying transmission rate (Brownian motion, B-splines, and approximate Gaussian processes). For these assessments, we use both simulated data and real-world data from SARS-CoV-2 in Geneva, Switzerland. We release the Bayesian workflow in an R package called HETTMO (“heterogeneous transmission model”), with the objective of promoting and facilitating access to this type of methods.

## Materials and methods

In accordance with best scientific practices and open science, we preregistered our methodology for this study on the Open Science Framework (OSF). This pre-registration document can be accessed at https://osf.io/n73gu/?view_only=4e469db4a58d428f99682e38c81f0d58.

### Transmission model

At its core, the compartmental infectious disease transmission model follows a Susceptible-Exposed-Infected-Removed (SEIR) structure (Figure 1). We extended this model by allowing the transmission rate to vary over time. The model definition is shown in Equation 1, where *ρ*(*t*) is the time-dependent factor that describes the change in the transmission rate over time relative to a baseline. The probability of a transmission event upon contact is *β*, *τ* is the inverse of the average incubation period, *γ* the inverse of the average infectious time and *c* is the average number of contacts an individual has per day. The total population is *N* = *S* + *E* + *I* + *R*. Both *τ* and *γ* are fixed, such that the generation time is 5.2 days, with this time equally distributed between the exposed and infected compartments [33]–[35]. Since our study period was less than a year, we could assume that there is no waning of immunity and that the total population size is constant.

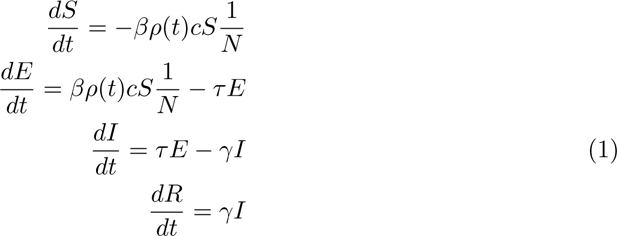

**Fig 1.**
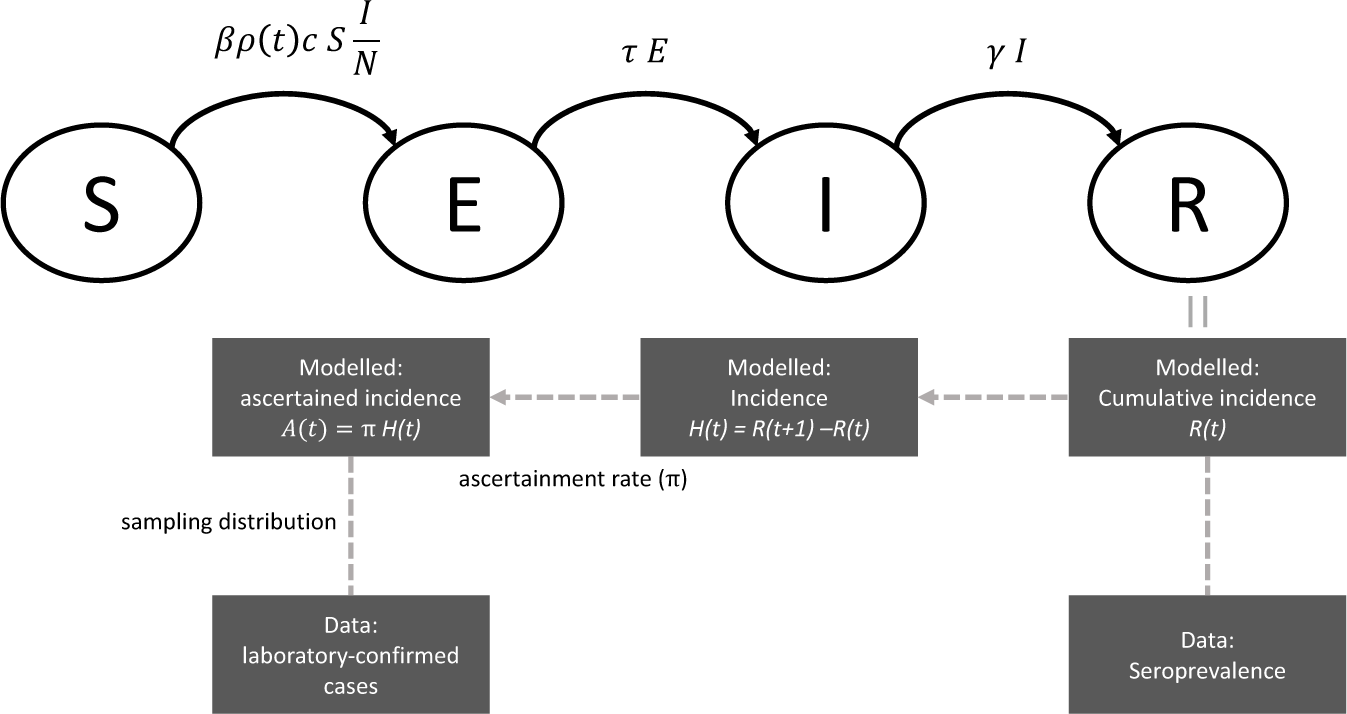
Schematic overview of the SEIR transmission model for SARS-CoV-2 and the steps to generate the number of laboratory confirmed cases and the observed seroprevalence.

### Feature 1: Adjustment for incomplete case ascertainment using seroprevalence data

From a given set of parameter values and initial conditions, the SEIR model generates the total number of newly recovered individuals in the population by unit of time (i.e. the true incidence by our definition, see Figure 1) as follows:

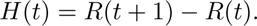

Only a fraction of this incidence will be ascertained as a laboratory-confirmed cases by testing positive (*A*(*t*)). The ascertainment rate *π* determines what fraction of the true incidence is observed: *A*(*t*) = *πH*(*t*). It is influenced by many determinants including testing activity and targeting, and may thus also vary over time. In this context the ascertainment rate is not statistically identifiable without the support of external data, such as a seroprevalence estimate. The seroprevalence is a measure of the number of recovered individuals in the population at a given time (if antibody waning and vaccination can be ignored), and thus informs about the cumulative true incidence over a period of time. Assuming that ascertainment is stable for that period of time, seroprevalence data can be used to estimate the ascertainment rate and anchor the model. In practice, we assume that the SEIR model also generates the cumulative number of removed individuals in the population at time *t* (from the *R* compartment), which is linked to seroprevalence data at time *t* using a simple binomial sampling distribution. We thus define periods bounded by seroprevalence studies, and estimate one ascertainment for each period. We also correct the seroprevalence data for imperfect testing [13].

### Feature 2: Sampling distribution for weekly laboratory-confirmed cases

We account for process noise in the transmission and observational noise in the ascertainment of cases by introducing a sampling distribution generating counts of laboratory-confirmed cases given the ascertained incidence. Process noise results from overdispersion of cases due to stochastic processes that are not captured by the compartmental transmission model, and observational noise from sampling of cases.

Here, we compare several options based on data from the Swiss canton of Geneva in 2020. First, we try a Poisson distribution, where the variance is equal to the mean *λ*. We then consider two distributions that include an additional overdispersion parameter *θ*: a quasi-Poisson model, where the variance is a linear function of the mean (*θλ*) and a negative binomial distribution, where the variance is a quadratic function of the mean (*λ* + *λ*^2^*θ*) [16].

### Feature 3: Flexible, time-varying transmission

In the compartmental transmission model, time-variation in transmission is controlled by the forcing function *ρ*(*t*), which applies to the contact rate *c* and the probability of transmission upon contact *β* at the same time. Therefore, time variation in these two components is considered together, and it is not possible to disentangle between them. We compare the performance and efficiency of three different methods to implement the time-varying transmission: Brownian motion, B-splines, and approximate Gaussian processes:

A We implemented Brownian motion as a Gaussian random walk similar to Bouranis et al (2022) with weekly time-steps; see Equations 2 and 3, taken from Bouranis et al (2022) [27]. In these equations, *t* is the discrete weekly time step, and *W* a random process whose elements are normally distributed with mean 0 and variance *s*. This value *s* is estimated from the data given a normal prior. This approach creates prior functions for *ρ*(*t*) with increasing variance over time [36]:

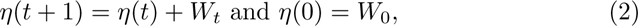

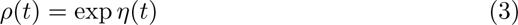

B Our implementation of the B-splines relies on the functions provided by Kharratzadeh [37]. B-splines are uniquely defined by the degree of the polynomials and the predefined set of knots. To be able to use the splines within the ODE system, without recomputing them every time the ODE is evaluated (that is, multiple times per MCMC iteration), we calculated the value of the B-splines for degree-1 points between two consecutive knots. Based on these values, we calculated the coefficients of a polynomial based on the degree of the B-spline using the Lagrange algorithm. These coefficients are then used as an input variable for the ODE model. For each iteration in the MCMC, a set of coefficients is sampled that defines how the B-splines must be combined to create the transmission rate function over time. In addition, this approach requires setting values for the knots. We consider five different sets of knots (Table 1) all in combination with cubic splines.

**Table 1.** Knot sequences.

| Knot sequence identifier | Location of first knot | Period between knots |
| --- | --- | --- |
| 1: true knots | 4 weeks | 4 weeks |
| 2: 8 weeks | 8 weeks | 8 weeks |
| 3: 12 weeks | 12 weeks | 12 weeks |
| 4: 4 weeks shifted | 6 weeks | 4 weeks |
| 5: 8 weeks shifted | 6 weeks | 8 weeks |

Overview of different sequences of knots used for the B-spline method to analyze time-dependent transmission rates in an compartmental infectious disease transmission model.

C Gaussian processes (GPs) are powerful and flexible fitting tools for modeling time series that are increasingly used in the field [38], [39]. We use a Gaussian process with an exponentiated quadratic covariance function, which, to our knowledge, has not yet been applied to compartmental transmission models. To reduce the computational cost, our implementation follows the proposition of Riutort-Mayol et al. (2020), using a basis function approximation via Laplace eigenfunctions, itself based on the mathematical theory developed by Solin and Säarkkäa (2020) [40], [41]. This low-rank Bayesian approximation requires several tuning parameters, most importantly the number of basis functions *M* and the boundary factor *c*, that determines the interval at which the approximation of the GP is valid. This interval is then given by the range of values at which the data is observed multiplied with the boundary factor. Both *M* and *c* influence the accuracy and the efficiency of the algorithm [40]. We test and compare the performance of the algorithm for a set of boundary factors and increasing number of basis-functions to find optimal values for the type of function we expect in our epidemiological data. Besides the number of basis functions and the boundary factor, the GP approximation also requires a parameter for the length scale (*L*, controlling the sinuosity of the basis-functions) and the marginal variance (*A*). As the length scale and the marginal variance both influence the smoothness of the function, they are unidentifiable in our set-up. We therefore fix the marginal variance to 0.5 and estimate the length scale from the data.

We note that both the Brownian motion and the B-splines are special cases of a Gaussian process given a specific kernel. However, our implementation of these methods differs from the implementation of the approximate Gaussian processes (aGPs).

### Feature 4: Stratification by age group

We consider three age groups in order to limit the computational cost: 0-19 years old, 20-64 years old and 65 and older. The stratification is implemented by replacing the contact rate *c* with a 3×3 contact matrix that indicates the average number of contacts an index case of a given age group (in the column) has with individuals of the other age groups (rows). We use a synthetic contact matrix as our pre-COVID baseline, as empirical data for Switzerland is lacking for this time period (Supplementary table 5). For this, we rely on the work of Prem et al. (2021) and rescale their suggested social contact matrix for Switzerland to match the age-distribution in our defined age groups in the canton of Geneva [42]. Stratification also applies to the processes of ascertainment, time-varying transmission and sampling that now occur independently by age group, hereby multiplying the number of parameters to estimate by three.

### Bayesian inference

We consider the models in a Bayesian framework, with the objective of estimating *β*, *ρ*(*t*), *π_t_*, and where relevant, *θ*, from two data sources: weekly counts of laboratory-confirmed cases of SARS-CoV-2 infection (this would also apply to any other respiratory virus) and one or more seroprevalence estimates. When relevant, these data need to be stratified by age group in the same way. We use weakly informative prior distributions for these parameters (Supplementary methods). The different versions of the model are implemented in Stan, a platform for Bayesian inference [8], [43]. Stan allows for coding a large variety of model features, relying on a few principles to optimize computational efficiency. For a detailed description of how to implement and scale-up ODE-based models, see Grinztajn et al. (2021) [11]. A key aspect here is the choice of the numerical ODE solver. To continue with our objective of identifying the most optimal implementation of the model in this type of situation, we compared all forward sensitivity solvers currently available in Stan: “rk45” (4th and 5th order Runge–Kutta-Fehlberg [44], [45]), “adams” (Adams-Moulton formula [46], [47]), “bdf” (backward differentiation formula [46], [47] and “ckrk” (fourth and fifth order explicit Runge-Kutta method for non-stiff and semi-stiff systems [45], [48]). We also compare to a simple solver that uses the trapezoidal rule to approximate the solution of the system as described in Bouranis et al [27]. For the trapezoidal solver, we use twenty equidistant time steps within each (weekly) time step in the model. Besides the solver itself, we also test different tuning values for the solver tolerance (1e-4, 1e-5 and 1e-6; not relevant for the trapezoidal solver) and the number of warmup-iterations (300 and 500). All combinations are run for 8 chains with 250 MCMC sampling-iterations.

### Simulated data

We validate and compare the different versions of the model with simulated data of an epidemic of a respiratory pathogen. The simulation study is conducted in two steps. In the first step, we assume that the population is well-mixed, and ignore the age stratification. We simulate data of laboratory-confirmed cases for 45 weeks and 100, 000 individuals, with two successive epidemic waves. We also simulate seroprevalence data after 20 weeks and at the end of the simulation, thus defining two periods with ascertainment *π*_1_ and *π*_2_, respectively. The transmission process is modeled with a probability of transmission per contact of *β* = 8.5%, a baseline of *c* = 11 contacts per day [42] and a time-varying component based on a spline of degree 3 and a knot every 4 weeks. We set ascertainment at *π*_1_ = 0.3 and *π*_2_ = 0.5. Supplementary Table 1 provides an overview of all parameter values chosen to create the simulated data. We generate one simulated dataset and apply all model versions to these data. We evaluate predictive performance by computing the root mean squared error between the estimated and true value of *ρ*(*t*), and evaluate computational performance by comparing the number of effective samples per second. In a second step, we select the best performing model version from step 1, add the stratification by age, and validate again on stratified simulated data. We modify several parameters to simulate a stratified dataset with three age classes (S6 Table). The simulated data are available in the HETTMO R package.

### Data from the canton of Geneva in 2020

Finally, we apply the best performing model version to weekly counts of laboratory-confirmed cases of SARS-CoV-2 infection in the canton of Geneva in 2020 (data from the Federal Office of Public Health). During this time period, we could reasonably assume that there was no waning of immunity and that vaccination did not yet influence transmission dynamics. Moreover, in Geneva, two seroprevalence surveys were performed during this time period: the first one from April 6th until May 10th [13], and the second from November 23th until December 23st [49]. The results are summarized in Table S8. Both surveys use the EuroImmune IgG test (Euroimmun; Lübeck, Germany #EI 2606-9601 G), which has a sensitivity of 93% and a specificity of 100% for the cutoff suggested by the manufacturer [50]. We aggregate all data according to the age groups used in the first serosurvey (0-19 years old, 20-64 years old and 65 and older). As the second serosurvey uses a different grouping, we reallocated the results in age group 18-24 to age groups 0-19 and 20-59 assuming a uniform distribution of age. The data for the canton of Geneva in 2020 are available in the HETTMO R-package.

### Software implementation

We use R version 4.2.1 [51]. We published a R package called HETTMO that contains all functions needed to perform the analysis and run the models. The package is based on Stan (version 2.21.7) [43] and the cmdstanr package (version 0.5.3) [52]. HETTMO is available on GitHub at https://github.com/JudithBouman2412/HETTMO.

## Results

We propose a Bayesian workflow for a compartmental transmission model aimed at analyzing the transmission of a respiratory virus in a population over a time period short enough so that immunity waning can be ignored (a few months or years). We focus on four aspects, representing four features deemed as essential in this situation. First, we validate in a simulation study that our models, jointly fitted to both laboratory-confirmed cases and seroprevalence data, are able to provide accurate and unbiased estimates of the ascertainment rate by periods of time bounded by serosurvey estimates (feature 1). We find that the appropriate handling of uncertainty in these models is largely influenced by the choice of sampling distributions (feature 2). We investigate the most adequate sampling distributions for laboratory-confirmed cases of SARS-CoV-2 using real data from the canton of Geneva (Switzerland). Whereas the Poisson and negative-binomial distribution under- and overestimates the variability in laboratory-confirmed cases, respectively, we found that the quasi-Poisson distribution, with the variance scaling linearly with the mean, better fits the variability of the data (Figure 2A).

**Fig 2.**
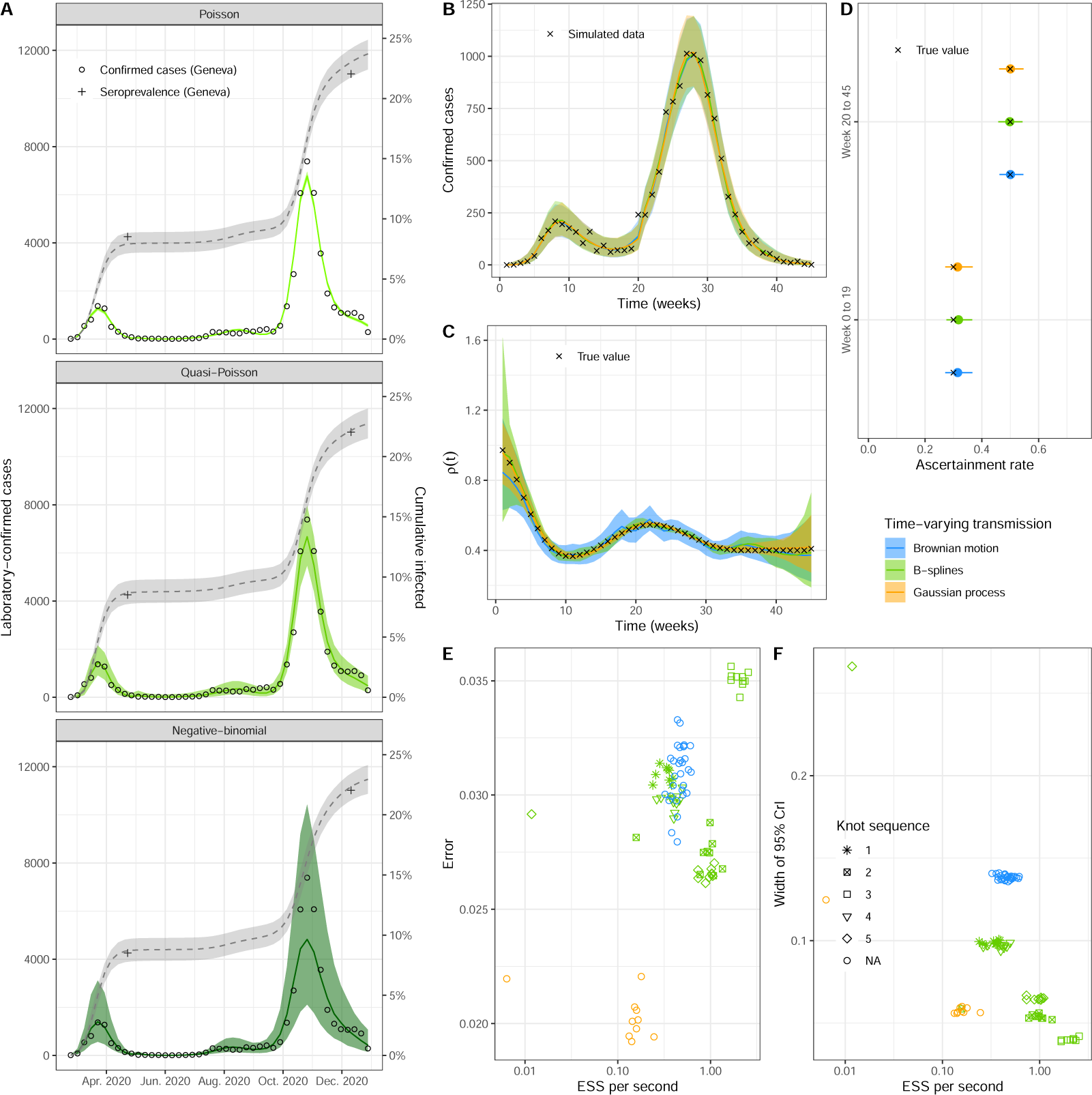
Result from unstratified models. (A) Posterior predictive plot for laboratory-confirmed cases (left y-axis, green ribbon) and cumulative incidence (right y-axis, gray ribbon) of SARS-CoV-2 in the canton of Geneva, Switzerland, for three iterations of the model with different sampling distributions (Poisson, quasi-Poisson and negative-binomial). Circles are weekly counts of laboratory-confirmed cases and pluses are estimates of seroprevalence at two time points. (B-D) Comparison of three methods of implementation of time-varying transmission on simulated data of a SARS-CoV-2 epidemic (posterior predictive plot, time-varying transmission *ρ*(*t*), and ascertainment rate by period). (E-F) Benchmark of different implementations of time-varying transmission on simulated data of a SARS-CoV-2 epidemic, with performance expressed in effective sample size (ESS) per second, error defined as the difference between the median posterior and true *ρ*(*t*), and the width of the 95% credible interval of *ρ*(*t*) as a measure for precision. See Table 1 1 for details about the knot sequence.

We then benchmark several implementations of the time-varying transmission in a simulation study (feature 3). These different approaches all use flexible parameterizations of forcing functions, estimated from data. In a systematic comparison, we confirm that implementations based on Brownian motion, B-splines, and aGPs lead to very similar model fits (Figure 2B). The estimation of the variation in transmission over time *ρ*(*t*) is accurate and unbiased under all three approaches (Figure 2C), but the Brownian motion approach overestimates the uncertainty. The estimation of the ascertainment rate *π_t_*is accurate under all three approaches, with a small overestimation in the first epidemic wave (Figure 2D).

Benchmarking the tuning parameters (the type of ODE solver, tolerance of the solver, and number of warm-up iterations) of the time-varying models in the simulation study highlights the importance of the ODE solver (Supplementary figure S1-S4). The choice of the solver can result, in some cases, in a factor of 1, 000 difference in performance (comparing Adams and trapezoidal solvers, S1 Fig). In terms of accuracy, the trapezoidal solver performs best for each time-varying model. However, with B-splines and aGPs, the average performance is increased by approximately 25%, with the ckrk solver. With aGPs, the performance also heavily depends on the choice of hyperparameters. Figure S3 shows the performance for a selection of number of basis functions and boundary factors. Across methods, 300 warm-up iterations appear sufficient for accurate model fits. For the comparison of the time-varying transmission rate models, we select the trapezoidal solver (Brownian motion) and the ckrk solver (B-splines and aGPs) and use 300 warm up iterations.

While leading to similar results, the three approaches differ in their computational performance measured by effective sample size (ESS) per second (Figures 2E-F). Depending on the pre-specified knot sequence, B-splines perform up to ten times faster than GPs in our example (average of GPs implementations compared to B-splines with knot sequence 3) and up to four times faster than Brownian motion (average of Brownian motion implementations compared to B-splines with knot sequence 3). While the error in the estimates is similar between B-splines and Brownian motion (Figure 2E), the width of the 95% credible interval of the estimation of *ρ*(*t*) is smaller for the Brownian motion model (Figure 2F). On the other hand, the width of the credible interval for *ρ*(*t*) was similar between aGPs and B-splines (Figure 2F), and the error in the estimate is smaller for aGPs compared to B-splines (Figure 2E). For our model and simulated data, B-splines perform best in terms of statistical accuracy and computational efficiency.

Building on the best performing model specification identified with non-stratified simulated data – ascertainment by period, quasi-Poisson sampling distribution, and B-spline implementation of time-varying transmission with the aforementioned tuning parameters – we consider the fourth feature of our model: age-stratification. Again, we first validate using simulated data of an epidemic of a respiratory infection with known parameters. In this case, both the laboratory-confirmed case data and the seroprevalence data are stratified in three age groups: 0-19 years, 20-64 years and 65+, modeling the interactions between these age groups with a synthetic contact matrix. We assume that each age group can have a different ascertainment rate per period. The model correctly captures the laboratory-confirmed cases as well as the seroprevalence data, and the estimates of the age- and time-specific ascertainment rates are accurate and precise (S5 Fig). The estimation of the time-variation in the transmission rate *ρ*(*t*) is in close correspondence with the true values when sufficient data are available. Larger deviations occur when the observed number of laboratory-confirmed cases is low.

In a last step, we apply the final iteration of the model, including all four essential features, to age-stratified laboratory-confirmed cases and seroprevalence data from the canton of Geneva, Switzerland, in 2020. The SARS-CoV-2 epidemic in Geneva in 2020, as in the rest of Switzerland, was characterized by a first wave in spring, low case counts in summer, followed by a severe second wave starting in the fall. Two serosurveys were conducted, once after each wave. The model is able to capture the dynamics of laboratory-confirmed cases and seroprevalence in each age group (Figure 3A). The dynamics of transmission as measured by *ρ*(*t*) are very similar across age groups during the first wave, but we observed a divergence from July 2020 onwards (Figure 3B).

**Fig 3.**
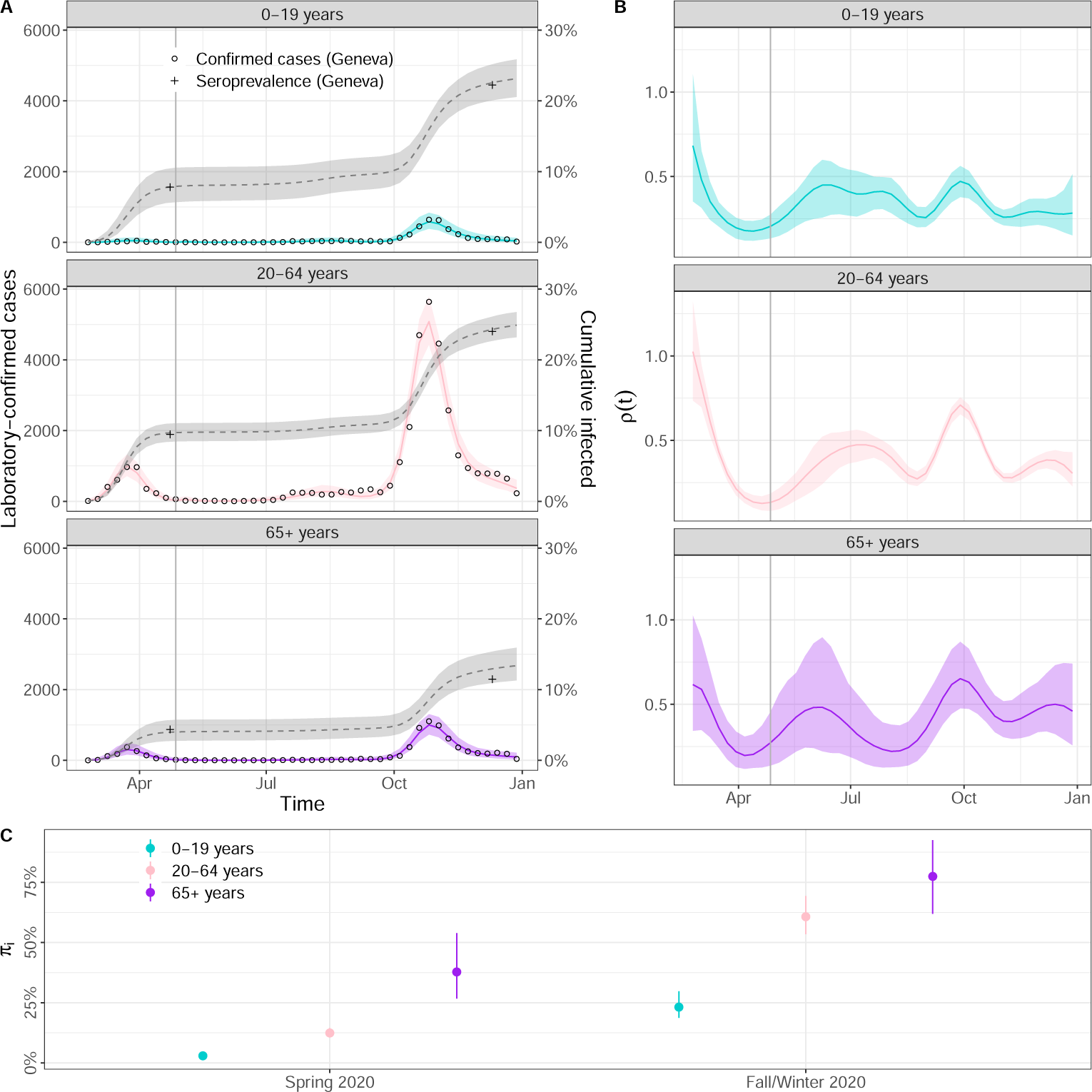
Modelled SARS-CoV-2 epidemic in Geneva, Switzerland, in 2020. (A) Posterior predictive plot for laboratory-confirmed cases (left y-axis, collored ribbon) and cumulative incidence (right y-axis, gray ribbon) per age group. Circles are weekly counts of laboratory-confirmed cases and pluses are estimates of seroprevalence at two time points. (B) Estimates of the time-varying change in transmission rate per age group using B-splines. (C) Estimates of the ascertainment rate per age group and time period.

Around this time, transmission decreased in the 65+ while remaining high all summer in the other age groups. There was then a temporary rise in transmission during fall that happened simultaneously in all age groups, but was the largest in magnitude in the 20-59 age group. Looking at ascertainment rates, we observe a clear improvement between spring and fall/winter, from 2.9% (95% CrI: 1.7 *−* 4.9%) to 23.2% (95% CrI: 18.0 *−* 31.1%) in age group 0-19, from 12.4% (95% CrI: 10.2 *−* 15.0%) to 60.7% (95% CrI: 52.4 *−* 70.9%) in age group 20-64 and from 37.8% (95% CrI: 25.1 *−* 58.1%) to 77.5% (95% CrI: 59.9 *−* 94.4%) in age group 65+.

## Discussion

Compartmental transmission models based on ODEs offer a principled and flexible way to study epidemics, but their implementation, handling, and computational efficiency for parameter inference using surveillance data can be challenging. With this study, we promote and facilitate access to methods that allow for reliable parameter inference, full propagation of uncertainty and integration of prior knowledge in compartmental transmission models. For this purpose, we developed a Bayesian workflow aimed at studying the spread of respiratory viruses such as SARS-CoV-2 in a population over a period of time where immunity waning can be ignored, with two commonly available data sources: laboratory-confirmed case and point estimates of seroprevalence. This workflow includes four main features deemed as essential for this task: adjustment for incomplete and differential case ascertainment across age groups, adequate sampling distribution, time-varying transmission rate and stratification by age. The exact implementation of two of these features, the sampling distribution and time-varying transmission, are the result of a benchmark and comparison of several methods using real and simulated data. We then apply this approach to real data on SARS-CoV-2 in the canton of Geneva in 2020. We also release the workflow as an out-of-the-box R package, where all model variations are available (https://github.com/JudithBouman2412/HETTMO).

Our work provides important methodological insights into fitting compartmental transmission models to surveillance data. First, we find that the variability in laboratory-confirmed case counts for SARS-CoV-2 was best described with a quasi-Poisson distribution, which is more commonly used in ecology to describe overdispersed data [16], [17]. The correct choice of this sampling distribution is critical for both inference and short-term forecasting of epidemic dynamics and can depend on the epidemiological situation. Further research could provide additional insights into how the process noise due to stochastic transmission and superspreading in combination with the observational noise from variations in testing results in this particular distribution. Second, we add to the existing literature on time-varying transmission rates for infectious diseases, by bringing empirical evidence that forcing functions based on B-splines appear to be the most effective way to implement flexible time-varying transmission in such models, with a clear advantage over Brownian motion and aGPs [25], [27]. Choosing good tuning parameters can increase performance by up to a factor of 1, 000 in some cases. Our comparison includes many different specifications regarding tuning parameters, allowing us to conclude with high confidence on this open question. Third, we demonstrate that a compartmental transmission model implemented in a Bayesian framework combined with MCMC is able to handle relatively high levels of complexity, with time-varying transmission and age-stratification, thereby highlighting its potential for future methodological developments.

The application of the model to the situation of the SARS-CoV-2 epidemic in the canton of Geneva, Switzerland, in 2020 highlights the practical advantages of our proposed approach. In the rather specific but critical situation of a newly-emerging respiratory pathogen circulating in a population, understanding the true level of transmission over time is of crucial importance to inform the public health response, but is generally concealed by the incomplete and unrepresentative ascertainment of cases. By combining information from laboratory-confirmed cases and serosurveys, our approach allows to estimate the ascertainment rate per age group by period bounded by seroprevalence estimates (or the emergence where seroprevalence is assumed to be null), and simultaneously to remove the effect of the ascertainment bias and determine the actual incidence of infection (with full uncertainty propagation). In the canton of Geneva, the overall ascertainment rate was estimated to be 8.6% during the first wave and 37% (95% CrI: 32 *−* 43%) during the second wave [13], [49]. These estimates from seroprevalence studies are somewhat lower than the across age group estimates from our model; (12.0% (95% CrI: 9.6 *−* 14.8%) and 54.7% (95% CrI: 47.4 *−* 64.1%)), respectively, because our second estimate includes all data since the end of the first serosurvey, whereas Stringhini et al. (2021) calculate ascertainment for the second wave based on data between September first and December 8th only. The large differences in age-specific case ascertainment during different periods of the pandemic highlight the importance of considering age-stratified models to monitor the epidemic dynamics of viral respiratory infections. Our estimates of the time-varying change in transmission allow us to compare variation in transmission due to changes in behavior, environment and NPIs across age groups while accounting for all other aspects included in the model (such as under-ascertainment and the accumulation of natural immunity). We found a consistent reduction in the transmission rate in all age groups after the implementation of strong NPIs in spring 2020. During summer 2020, the relative transmission in 65+-year-olds was somewhat lower compared to the other age groups which could be a result of more careful social contact behavior as reported laboratory-confirmed cases numbers started to increase. At the beginning of the second wave in fall 2020, the comparatively higher transmission in 20-64 year olds compared to 0-19-year-olds is in favor of an epidemic relapse that can be attributed more to working people.

This work also has a number of limitations. First, the benchmark results are specific to our simulated data and our choice of prior distributions. We chose weakly-specific priors, only limiting the range of possible observations to plausible values [53], [54]. A prior predictive check is shown in Supplementary Figure S5. Second, it is possible that the relative performance of the three presented methods differs for distinct datasets, for instance for data collected during a longer time-period, different epidemic dynamics or a different infectious disease. The publication of our workflow in the HETTMO R package allows users to apply all presented methods and evaluate which one is the most suitable for their data. Third, for the approximate Gaussian process model, performance depends heavily on the choice of the hyperparameters (S3 Fig).

Additionally, when applying the B-spline based method from this package, one should be aware that the performance of this method depends on the choice of the knots (Figure 2). A sufficient number of knots can be identified by subsequently increasing their number until the estimate of the transmission rate does not change any more. Fourth, our benchmark focuses on the ability of the different methods to estimate the time-varying transmission rate during the period for which data were available. We did not compare the precision for short-term forecasting. However, based on the characteristics of the methods, we would advise to use either the Brownian motion or aGP model for prediction, because for these method the variance increases with time. In contrast, B-splines are known to be at risk of error in extrapolations. Fifth, the current version of HETTMO is only useful in a limited range of situations, i.e. in a relatively short period of time following the emergence of a respiratory virus, in order to fulfill different assumptions (entirely susceptible population at the start, no vaccination, no waning of immunity and negligible changes in population sizes). However, it provides a starting point for extensions relaxing these assumptions.

While enormous amounts of data have been generated during the early stages of the SARS-CoV-2 pandemic, the complexity involved, with differential under-ascertainment, transmission and immunity all varying in time, creates various challenges in their interpretation. Approaches from the field of infectious disease modeling can bring invaluable insights in situations of epidemics, but require adequate, validated and efficient tools. By combining the structure of ODE based compartmental transmission models and the power of full Bayesian inference, HETTMO provides such a tool, in the form of a Bayesian workflow, for relatively simple situations: a newly-emerging respiratory virus spreading in a population before vaccination and immunity waning can play a role. While individual features of our workflow have been described in the literature, prior studies have not conducted a comprehensive comparison of various implementation methods to develop a complete Bayesian workflow for this specific problem. The further development of infectious disease models that can be fitted to various data sources in a Bayesian framework will promote their use for real-time monitoring, short-term forecasting, and policy making.

## Supplementary methods

### Definition of priors

We chose a weakly-informative gamma prior on the initial basic reproduction number (*R*_0_) of the model, where we set the mean and variance to 2.5 (shape =2.5 and scale = 1, corresponding to an expected *R*_0_ between 0.4 and 6.4 based on the 2.5% and 97.5% percentiles). This basic reproduction number then defines the initial probability of transmission by the relationship derived from the ODE system: 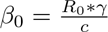, where *c* and *γ* are assumed to be fixed and known. For the initial number of infected individuals (*I*(0)), we also took a gamma distribution (mean = 0.25, variance=0.25). For the ascertainment rate, we took a beta(2,2) distribution. The prior for the overdispersion parameter of the quasi-Poisson model (*θ*) is exponential distribution with mean 0.1. For the negative binomial model, we use a exponential distribution with *λ* = 1 as the prior on the inverse of the dispersion parameter (*ϕ*).

### Priors specific for Brownian motion model

The prior on the parameters specific for the Brownian motion model are defined such that the weekly transmission rate 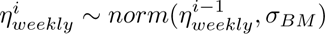. Where the prior on BM is a normal distribution with a zero mean and standard deviation equal to 0.1.

### Priors specific for B-splines model

For the spline model, we took a gamma prior with shape equal to 2.5 and scale equal to 5, resulting in 0.083 and 1.28 as the 2.5% and 97.5% percentiles.

### Priors specific for approximate Gaussian processes model

The regression weights are standard normally distributed. The prior on the length scale parameter is a normal distribution with mean equal to 0 and the standard deviation of 3 and for the marginal variance we use an exponential distribution with a rate parameter of 5.

## Data Availability

Data and code are available as an R-package on https://github.com/JudithBouman2412/HETTMO.

## Acknowledgments

We warmly thank Andrew Gelman, Seth Flaxman, Elizaveta Semenova, Samir Bhatt, and David Ginsbourger for helpful advice at various stages of this project.

---

**S1 Fig.**
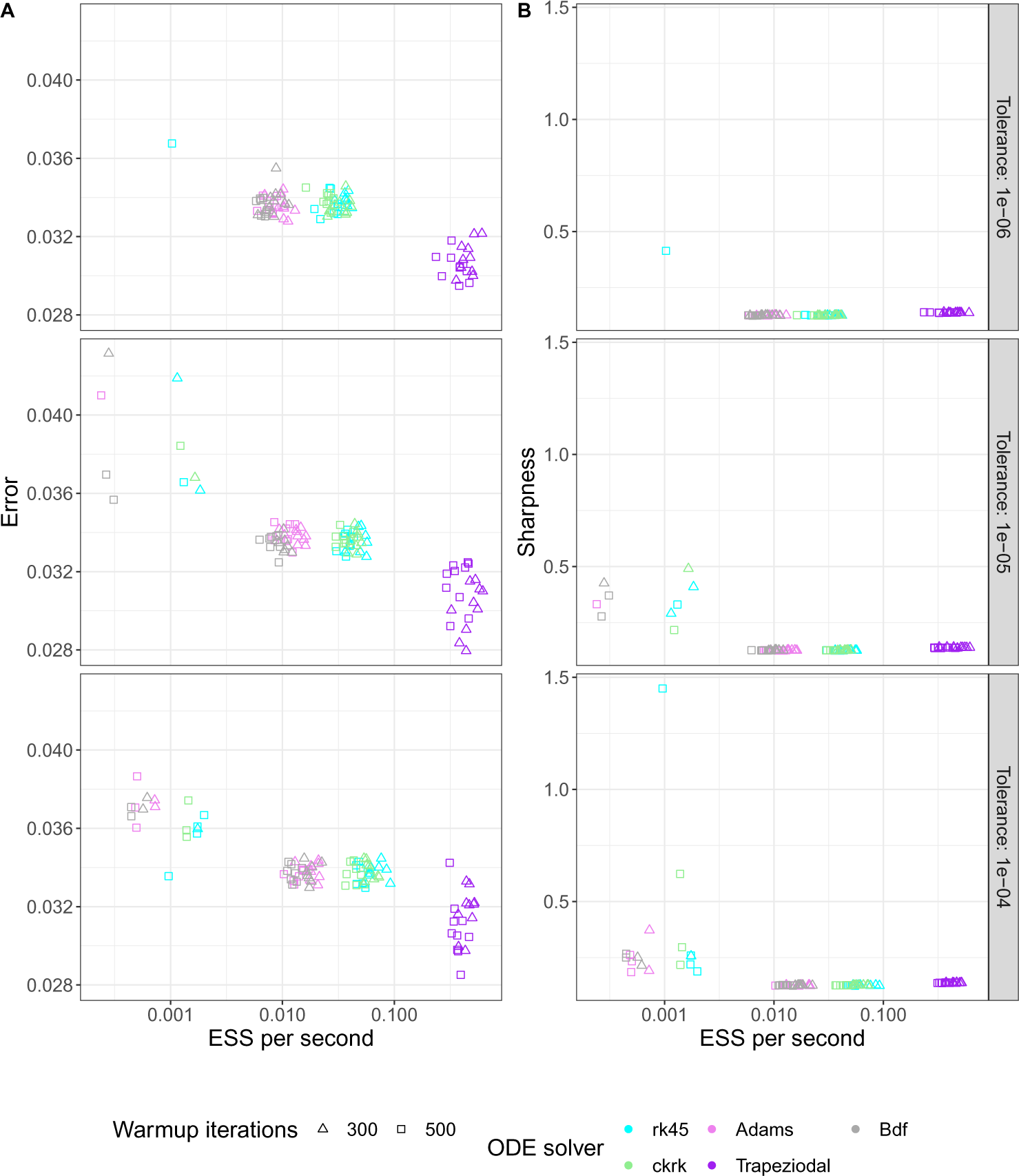
Benchmark for Brownian motion model. Comparison of computational performance for the Brownian motion model of the time-varying transmission rate of SARS-CoV-2 for simulated, non-stratified data for various tuning parameters: tolerance, ODE solver and number of warmup iterations. (A) The root mean square error (RMSE) in estimating the time-variation in the transmission. (B) The sharpness (size of the 90% confidence interval) of the time-variation in the transmission.

**S2 Fig.**
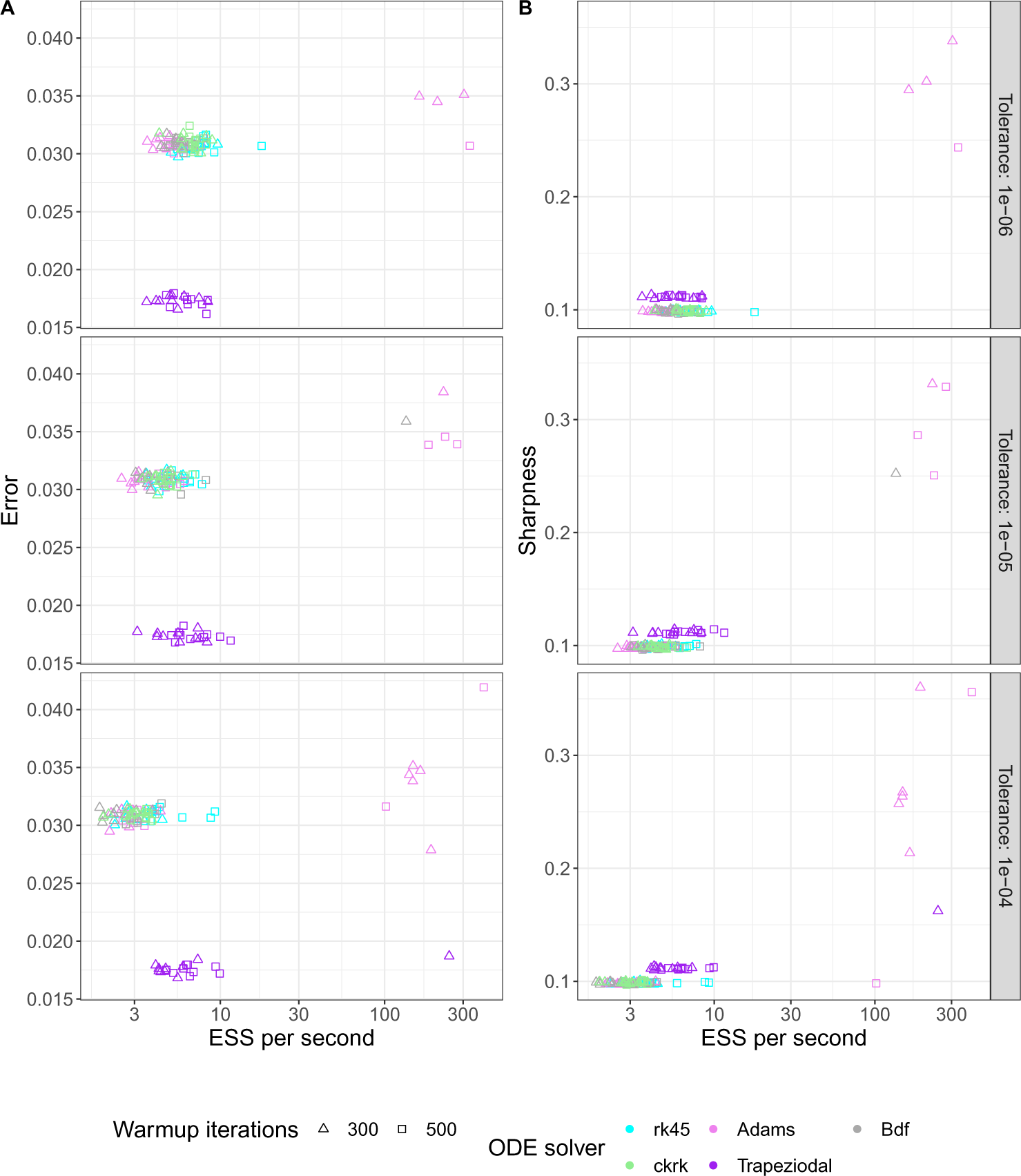
Benchmark for B-spline model. Comparison of computational performance for the B-spline model of the time-varying transmission rate of SARS-CoV-2 for simulated, non-stratified data for various tuning parameters: tolerance, ODE solver and number of warmup iterations. (A) The root mean squared error (RMSE) in estimating the time-variation in the transmission. (B) The sharpness (size of the 90% confidence interval) of the time-variation in the transmission.

**S3 Fig.**
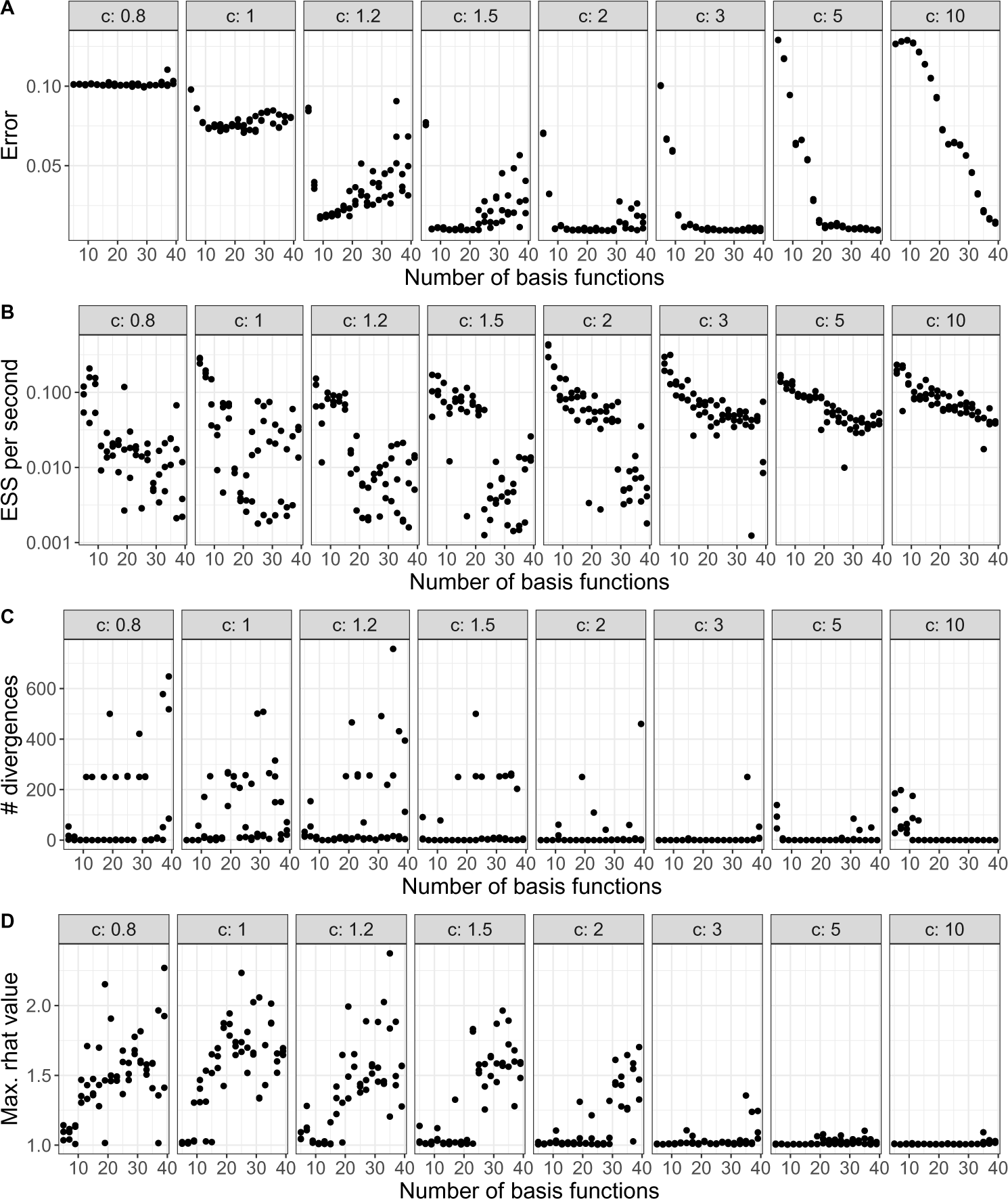
Benchmark for hyper-parameters approximate Gaussian processes model. Analysis of the optimal number of basis functions and boundary factor for the approximate Gaussian Processes based time-varying transmission model of SARS-CoV-2 using simulated data. The number of warm up and sampling iterations are both fixed to 300 and the trapezoidal solver is used.

**S4 Fig.**
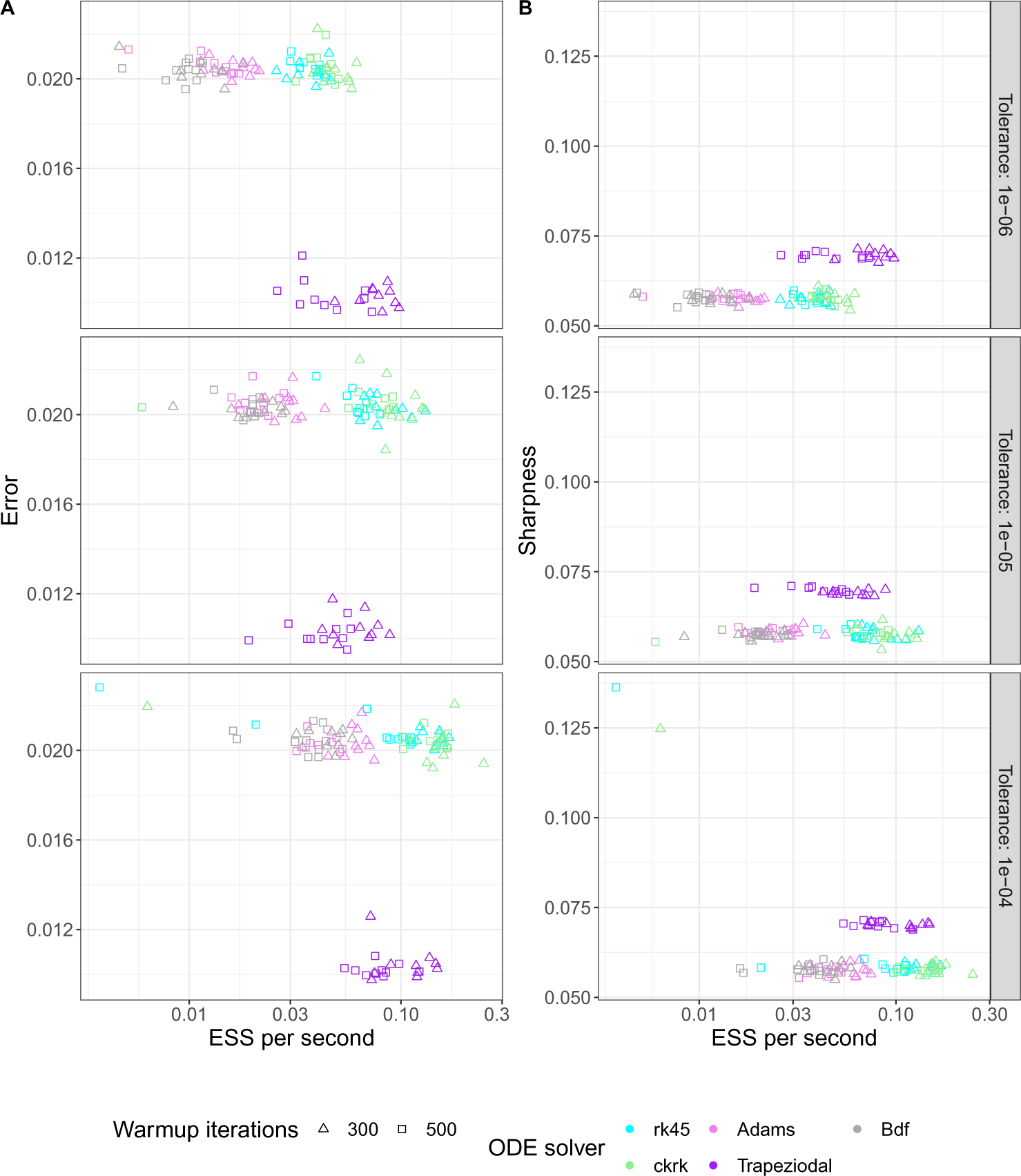
Benchmark for approximate Gaussian processes model. Comparison of computational performance for the approximate Gaussian processes model of the time-varying transmission rate of SARS-CoV-2 for simulated, non-stratified data for various tuning parameters: tolerance, ODE solver and number of warmup iterations. (A) The root mean squared error (RMSE) in estimating the time-variation in the transmission. (B) The sharpness (size of the 90% confidence interval) of the time-variation in the transmission.

**S5 Fig.**
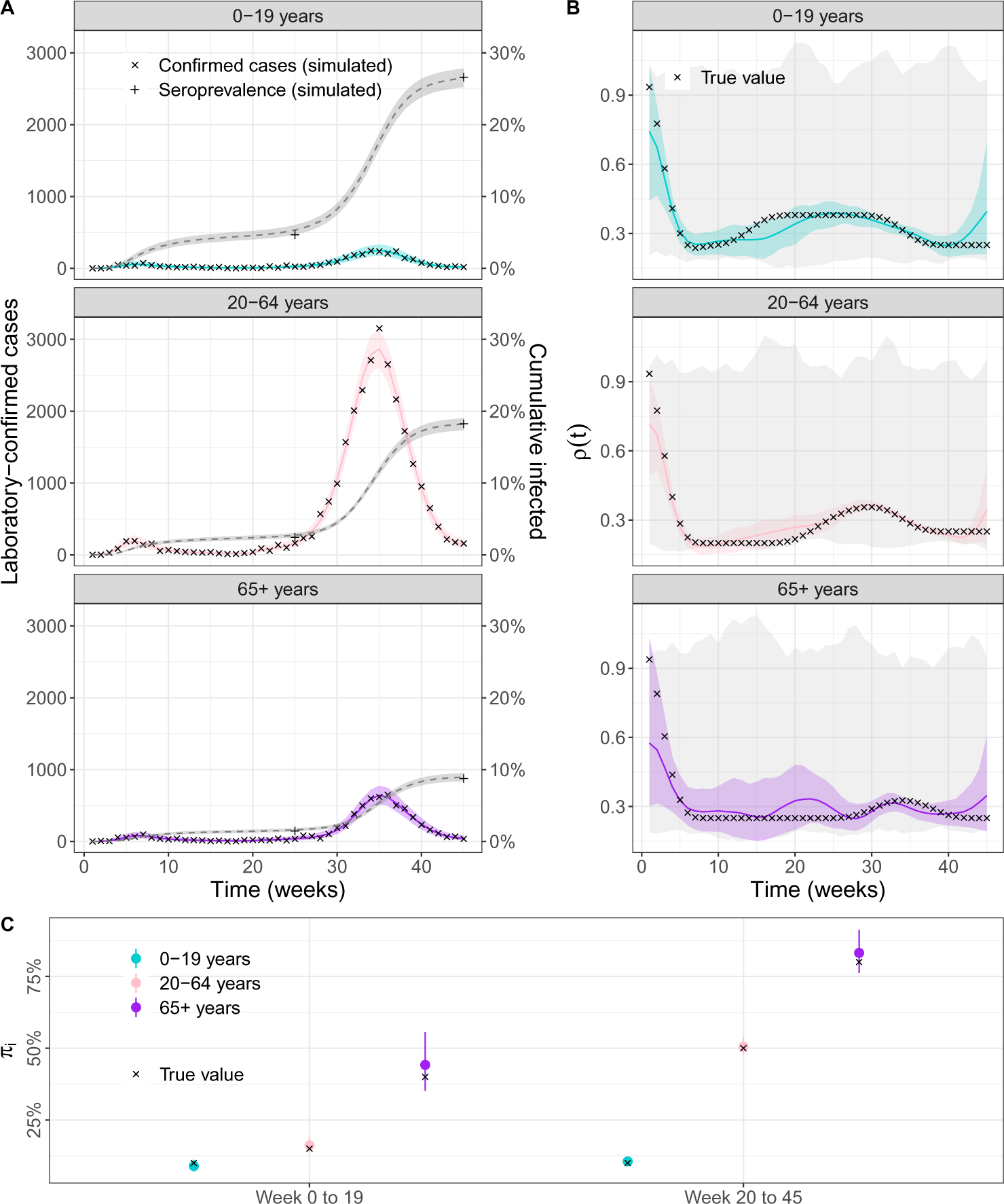
Model results for stratified simulated data. (A) Posterior predictive plot for laboratory-confirmed cases (left y-axis, collored ribbon) and cumulative incidence (right y-axis, gray ribbon) per age-group using the B-spline based age-stratified model applied to simulated data. Crosses are weekly simulated counts of laboratory-confirmed cases and pluses are simulated estimates of seroprevalence at two time points. (B) Estimates of the time-varying change in transmission rate per age group using B-splines. Crosses represent the true, simulated values. (C) Estimates of the ascertainment rate per age group and time period. Crosses represent the true, simulated values.

**S6 Table:** Parameters for simulating unstratified data using the B-splines model.

| Parameter | Value | Explanation | Estimated/fixed |
| --- | --- | --- | --- |
| $N$ | 100,000 | Total population size | Fixed |
| $S(0)$ | 0 | Initial value | Fixed |
| $E(0)$ | 0 | Initial value | Fixed |
| $I(0)$ | 3 | Initial value | Fixed |
| $R(0)$ | 0 | Initial value | Fixed |
| $\beta$ | 8.5% | Probability of infection upon contact [55] | Fixed |
| $c$ | 11 per day | Contact rate [42] | Fixed |
| $\gamma$ | 2.7 days | Average stay in compartment E [33]–[35] | Fixed |
| $\tau$ | 2.7 days | Average stay in compartment I [33]–[35] | Fixed |
| $\pi_1$ | 30% | Ascertainment rate during first time period | Estimated |
| $\pi_2$ | 50% | Ascertainment rate during second time period | Estimated |
| $t_1$ | 20 weeks | Time of transition between time period 1 and 2 | Fixed |
| $\theta$ | 5 | Overdispersion parameter of quasi-Poisson distribution | Estimated |
| $t$ | 45 weeks | Total duration of simulation | Fixed |
| $N_s$ | 5,000 | Number of seroprevalence tests simulated | Fixed |

**S7 Table:**
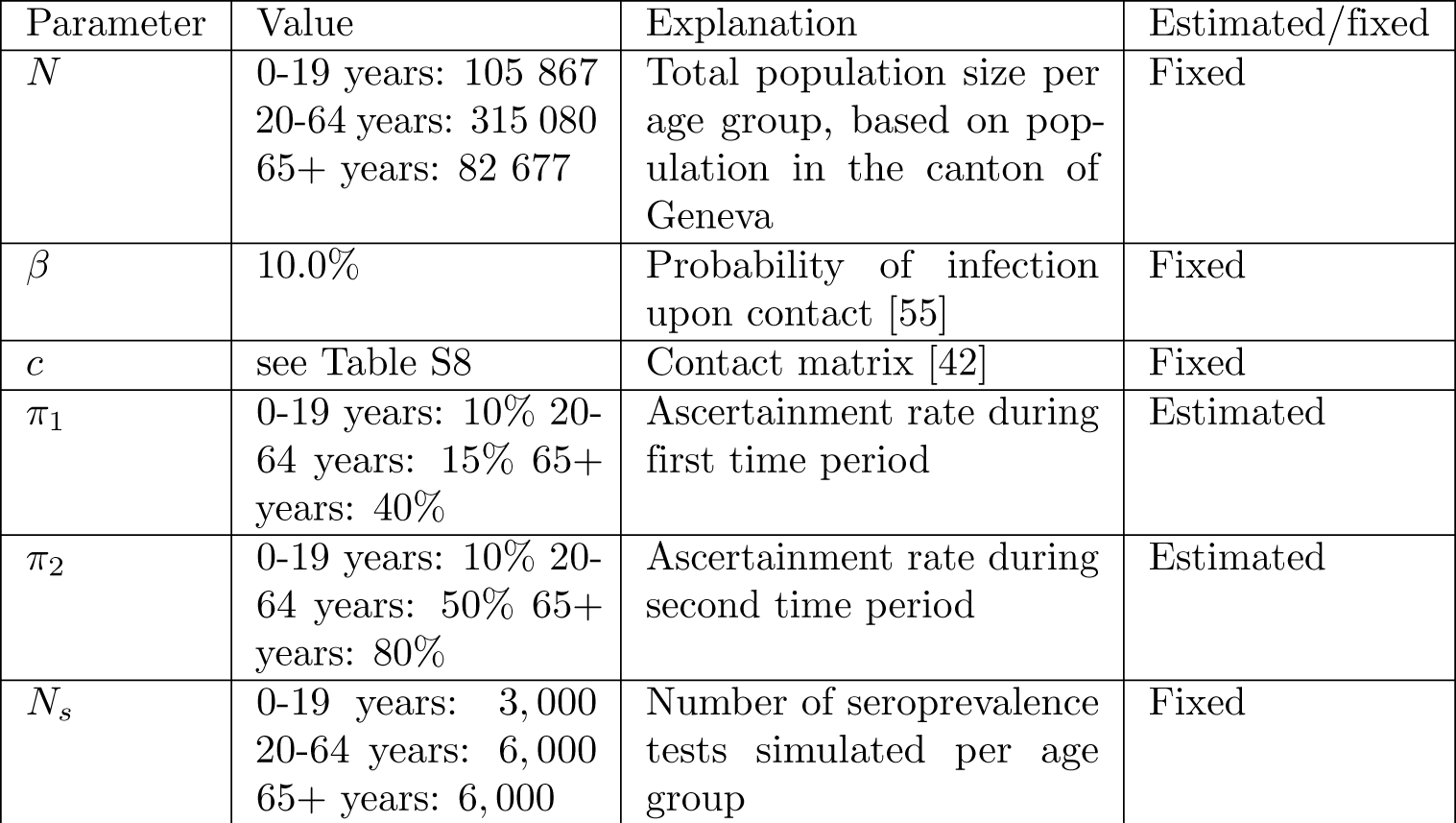
Parameters adapted from the unstratified model for simulating stratified data using the B-splines.

| Parameter | Value | Explanation | Estimated/fixed |
| --- | --- | --- | --- |
| $N$ | 0-19 years: 105 867<br>20-64 years: 315 080<br>65+ years: 82 677 | Total population size per age group, based on population in the canton of Geneva | Fixed |
| $\beta$ | 10.0% | Probability of infection upon contact [55] | Fixed |
| $c$ | see Table S8 | Contact matrix [42] | Fixed |
| $\pi_1$ | 0-19 years: 10% 20-64 years: 15% 65+ years: 40% | Ascertainment rate during first time period | Estimated |
| $\pi_2$ | 0-19 years: 10% 20-64 years: 50% 65+ years: 80% | Ascertainment rate during second time period | Estimated |
| $N_s$ | 0-19 years: 3,000<br>20-64 years: 6,000<br>65+ years: 6,000 | Number of seroprevalence tests simulated per age group | Fixed |

**S8 Table:** Seroprevalence data from the canton of Geneva, obtained from Stringhini et al (2020) and Stringhini et al (2021) [13], [49].

| Age group | Number of tests performed, spring 2020 | Number of positive tests, spring 2020 | Number of tests performed, fall/winter 2020 | Number of positive tests, fall/winter 2020 |
| --- | --- | --- | --- | --- |
| 0-19 years | 455 | 33 | 1083 | 224 |
| 20-64 years | 1942 | 171 | 2439 | 545 |
| 65+ years | 369 | 15 | 478 | 51 |

**S9 Table:** Contact matrix indicating the average number of contacts per day of an index case (column) with individuals in three defined age groups. This matrix was constructed using Prem et al (2021) and is adjusted for the population structure of the canton of Geneva [42]. This matrix is used both for simulating stratified SARS-CoV-2 data and for analysing the data from the canton of Geneva.

| Age of contact: |  |  |  |
| --- | --- | --- | --- |
| 0-19 years | 9.3 | 6.9 | 0.9 |
| 20-64 years | 2.2 | 10.6 | 1.2 |
| 65+ years | 1.0 | 4.4 | 1.8 |
| Age of index case: | 0-19 years | 20-64 years | 65+ years |

## References

[1] W. O. Kermack and A. G. McKendrick, “A contribution to the mathematical theory of epidemics,” Proceedings of the royal society of london. Series A, Containing papers of a mathematical and physical character, vol. 115, no. 772, pp. 700–721, 1927.

[2] J. Arino, F. Brauer, P. Van Den Driessche, J. Watmough, and J. Wu, “A model for influenza with vaccination and antiviral treatment,” Journal of theoretical biology, vol. 253, no. 1, pp. 118–130, 2008.

[3] N. M. Ferguson, D. A. Cummings, C. Fraser, J. C. Cajka, P. C. Cooley, and D. S. Burke, “Strategies for mitigating an influenza pandemic,” Nature, vol. 442, no. 7101, pp. 448–452, 2006.

[4] C. Viboud, O. N. Bjørnstad, D. L. Smith, L. Simonsen, M. A. Miller, and B. T. Grenfell, “Synchrony, waves, and spatial hierarchies in the spread of influenza,” science, vol. 312, no. 5772, pp. 447–451, 2006.

[5] M. J. Keeling, E. M. Hill, E. E. Gorsich, et al., “Predictions of covid-19 dynamics in the uk: Short-term forecasting and analysis of potential exit strategies,” PLoS computational biology, vol. 17, no. 1, e1008619, 2021.

[6] B. Tepekule, A. Hauser, V. N. Kachalov, et al., “Assessing the potential impact of transmission during prolonged viral shedding on the effect of lockdown relaxation on covid-19,” PLoS computational biology, vol. 17, no. 1, e1008609, 2021.

[7] M. D. Hoffman, A. Gelman, et al., “The no-u-turn sampler: Adaptively setting path lengths in hamiltonian monte carlo.,” J. Mach. Learn. Res., vol. 15, no. 1, pp. 1593–1623, 2014.

[8] B. Carpenter, A. Gelman, M. D. Hoffman, et al., “Stan: A probabilistic programming language,” Journal of statistical software, vol. 76, 2017.

[9] D. Phan, N. Pradhan, and M. Jankowiak, “Composable effects for flexible and accelerated probabilistic programming in numpyro,” arXiv preprint arXiv:1912.11554, 2019.

[10] H. Ge, K. Xu, and Z. Ghahramani, “Turing: A language for flexible probabilistic inference,” pp. 1682–1690, 2018. [Online]. Available: http://proceedings.mlr.press/v84/ge18b.html.

[11] L. Grinsztajn, E. Semenova, C. C. Margossian, and J. Riou, “Bayesian workflow for disease transmission modeling in stan,” Statistics in medicine, vol. 40, no. 27, pp. 6209–6234, 2021.

12. A. Gelman, A. Vehtari, D. Simpson, et al., “Bayesian workflow,” arXiv preprint arXiv:2011.01808, 2020.

[13] S. Stringhini, A. Wisniak, G. Piumatti, et al., “Seroprevalence of anti-sars-cov-2 igg antibodies in geneva, switzerland (serocov-pop): A population-based study,” The Lancet, vol. 396, no. 10247, pp. 313–319, 2020.

[14] J. Perez-Saez, S. A. Lauer, L. Kaiser, et al., “Serology-informed estimates of sars-cov-2 infection fatality risk in geneva, switzerland,” The Lancet Infectious Diseases, vol. 21, no. 4, e69–e70, 2021.

[15] T. W. Russell, N. Golding, J. Hellewell, et al., “Reconstructing the early global dynamics of under-ascertained covid-19 cases and infections,” BMC medicine, vol. 18, no. 1, p. 332, 2020.

[16] J. M. Ver Hoef and P. L. Boveng, “Quasi-poisson vs. negative binomial regression: How should we model overdispersed count data?” Ecology, vol. 88, no. 11, pp. 2766–2772, 2007.

[17] A. Lindén and S. Mäntyniemi, “Using the negative binomial distribution to model overdispersion in ecological count data,” Ecology, vol. 92, no. 7, pp. 1414–1421, 2011.

[18] G. Chowell, N. W. Hengartner, C. Castillo-Chavez, P. W. Fenimore, and J. M. Hyman, “The basic reproductive number of ebola and the effects of public health measures: The cases of congo and uganda,” Journal of theoretical biology, vol. 229, no. 1, pp. 119–126, 2004.

[19] J. P. Chávez, T. Gäotz, S. Siegmund, and K. P. Wijaya, “An sir-dengue transmission model with seasonal effects and impulsive control,” Mathematical biosciences, vol. 289, pp. 29–39, 2017.

[20] A. Hauser, M. J. Counotte, C. C. Margossian, et al., “Estimation of sars-cov-2 mortality during the early stages of an epidemic: A modeling study in hubei, china, and six regions in europe,” PLoS medicine, vol. 17, no. 7, e1003189, 2020.

[21] J. Brugger and C. L. Althaus, “Transmission of and susceptibility to seasonal influenza in switzerland from 2003 to 2015,” Epidemics, vol. 30, p. 100 373, 2020.

[22] D. He, J. Dushoff, T. Day, J. Ma, and D. J. Earn, “Mechanistic modelling of the three waves of the 1918 influenza pandemic,” Theoretical Ecology, vol. 4, pp. 283–288, 2011.

[23] H. G. Hong and Y. Li, “Estimation of time-varying reproduction numbers underlying epidemiological processes: A new statistical tool for the covid-19 pandemic,” PloS one, vol. 15, no. 7, e0236464, 2020.

[24] H. Inouzhe, M. X. Rodriguez-Alvarez, L. Nagar, and E. Akhmatskaya, “Dynamic sir/seir-like models comprising a time-dependent transmission rate: Hamiltonian monte carlo approach with applications to covid-19,” arXiv preprint arXiv:2301.06385, 2023.

[25] B. Cazelles, C. Champagne, and J. Dureau, “Accounting for non-stationarity in epidemiology by embedding time-varying parameters in stochastic models,” PLoS computational biology, vol. 14, no. 8, e1006211, 2018.

[26] B. Cazelles, C. Champagne, B. Nguyen-Van-Yen, C. Comiskey, E. Vergu, and B. Roche, “A mechanistic and data-driven reconstruction of the time-varying reproduction number: Application to the covid-19 epidemic,” PLoS computational biology, vol. 17, no. 7, e1009211, 2021.

[27] L. Bouranis, N. Demiris, K. Kalogeropoulos, and I. Ntzoufras, “Bayesian analysis of diffusion-driven multi-type epidemic models with application to covid-19,” arXiv preprint arXiv:2211.15229, 2022.

[28] L. Pellis, S. Cauchemez, N. M. Ferguson, and C. Fraser, “Systematic selection between age and household structure for models aimed at emerging epidemic predictions,” Nature communications, vol. 11, no. 1, p. 906, 2020.

[29] S. Ranjeva, R. Subramanian, V. J. Fang, et al., “Age-specific differences in the dynamics of protective immunity to influenza,” Nature Communications, vol. 10, no. 1, p. 1660, 2019.

[30] N. G. Davies, P. Klepac, Y. Liu, K. Prem, M. Jit, and R. M. Eggo, “Age-dependent effects in the transmission and control of covid-19 epidemics,” Nature medicine, vol. 26, no. 8, pp. 1205–1211, 2020.

[31] A. T. Levin, W. P. Hanage, N. Owusu-Boaitey, K. B. Cochran, S. P. Walsh, and G. Meyerowitz-Katz, “Assessing the age specificity of infection fatality rates for covid-19: Systematic review, meta-analysis, and public policy implications,” European journal of epidemiology, vol. 35, no. 12, pp. 1123–1138, 2020.

[32] B. Wachtler, N. Michalski, E. Nowossadeck, et al., “Socioeconomic inequalities in the risk of sars-cov-2 infection–first results from an analysis of surveillance data from germany,” Journal of Health Monitoring, vol. 5, no. Suppl 7, p. 18, 2020.

[33] T. Ganyani, C. Kremer, D. Chen, et al., “Estimating the generation interval for coronavirus disease (covid-19) based on symptom onset data, march 2020,” Eurosurveillance, vol. 25, no. 17, p. 2000257, 2020.

[34] M. Alene, L. Yismaw, M. A. Assemie, D. B. Ketema, W. Gietaneh, and T. Y. Birhan, “Serial interval and incubation period of covid-19: A systematic review and meta-analysis,” BMC Infectious Diseases, vol. 21, no. 1, pp. 1–9, 2021.

[35] W. S. Hart, S. Abbott, A. Endo, et al., “Inference of the sars-cov-2 generation time using uk household data,” Elife, vol. 11, e70767, 2022.

[36] B. Øksendal, “Stochastic differential equations,” in Stochastic differential equations, Springer, 2003, pp. 65–84.

[37] M. Kharratzadeh, “Splines in stan,” mc-stan documentation, 2017. [Online]. Available: https://mc-stan.org/users/documentation/case-studies/splines_in_stan.html.

[38] L. M. G. Rincon, E. M. Hill, L. Dyson, M. J. Tildesley, and M. J. Keeling, “Bayesian estimation of real-time epidemic growth rates using gaussian processes: Local dynamics of sars-cov-2 in england (preprint),” 2022.

[39] S. Abbott, J. Hellewell, K Sherratt, et al., “Epinow2: Estimate real-time case counts and time-varying epidemiological parameters,” R package version 0.1.0, 2020.

40. G. Riutort-Mayol, P.-C. Bürkner, M. R. Andersen, A. Solin, and A. Vehtari, “Practical hilbert space approximate bayesian gaussian processes for probabilistic programming,” arXiv preprint arXiv:2004.11408, 2020.

[41] A. Solin and S. Särkkä, “Hilbert space methods for reduced-rank gaussian process regression,” Statistics and Computing, vol. 30, no. 2, pp. 419–446, 2020.

[42] K. Prem, K. v. Zandvoort, P. Klepac, et al., “Projecting contact matrices in 177 geographical regions: An update and comparison with empirical data for the covid-19 era,” PLoS computational biology, vol. 17, no. 7, e1009098, 2021.

[43] Stan Development Team, “Stan modeling language users guide and reference manual,” vol. version 2.30, 2022. [Online]. Available: https://mc-stan.org.

[44] J. R. Dormand and P. J. Prince, “A family of embedded runge-kutta formulae,” Journal of computational and applied mathematics, vol. 6, no. 1, pp. 19–26, 1980.

[45] K. Ahnert and M. Mulansky, “Odeint–solving ordinary differential equations in c++,” in AIP Conference Proceedings, American Institute of Physics, vol. 1389, 2011, pp. 1586–1589.

[46] S. D. Cohen, A. C. Hindmarsh, and P. F. Dubois, “Cvode, a stiff/nonstiff ode solver in c,” Computers in physics, vol. 10, no. 2, pp. 138–143, 1996.

[47] R. Serban and A. C. Hindmarsh, “Cvodes: The sensitivity-enabled ode solver in sundials,” in International Design Engineering Technical Conferences and Computers and Information in Engineering Conference, vol. 47438, 2005, pp. 257–269.

[48] F. Mazzia, J. R. Cash, and K. Soetaert, “A test set for stiff initial value problem solvers in the open source software r: Package detestset,” Journal of Computational and Applied Mathematics, vol. 236, no. 16, pp. 4119–4131, 2012.

[49] S. Stringhini, M.-E. Zaballa, J. Perez-Saez, et al., “Seroprevalence of anti-sars-cov-2 antibodies after the second pandemic peak,” The Lancet Infectious Diseases, vol. 21, no. 5, pp. 600–601, 2021.

[50] B. Meyer, G. Torriani, S. Yerly, et al., “Validation of a commercially available sars-cov-2 serological immunoassay,” Clinical microbiology and infection, vol. 26, no. 10, pp. 1386–1394, 2020.

[51] R Core Team, R: A language and environment for statistical computing, R Foundation for Statistical Computing, Vienna, Austria, 2022. [Online]. Available: https://www.R-project.org/.

[52] J. Gabry and R. Češnovar, Cmdstanr: R interface to ‘cmdstan’, https://mc-stan.org/cmdstanr/, https://discourse.mc-stan.org, 2022.

[53] A. Gelman, A. Jakulin, M. G. Pittau, and Y.-S. Su, “A weakly informative default prior distribution for logistic and other regression models,” 2008.

[54] J. Gabry, D. Simpson, A. Vehtari, M. Betancourt, and A. Gelman, “Visualization in bayesian workflow,” Journal of the Royal Statistical Society Series A: Statistics in Society, vol. 182, no. 2, pp. 389–402, 2019.

[55] C. I. Jarvis, K. Van Zandvoort, A. Gimma, et al., “Quantifying the impact of physical distance measures on the transmission of covid-19 in the uk,” BMC medicine, vol. 18, no. 1, pp. 1–10, 2020.

